# ChatGPT for assessing risk of bias of randomized trials using the RoB 2.0 tool: A methods study

**DOI:** 10.1101/2023.11.19.23298727

**Authors:** Tyler Pitre, Tanvir Jassal, Jhalok Ronjan Talukdar, Mahnoor Shahab, Michael Ling, Dena Zeraatkar

## Abstract

**Background:** Internationally accepted standards for systematic reviews necessitate assessment of the risk of bias of primary studies. Assessing risk of bias, however, can be time- and resource-intensive. AI-based solutions may increase efficiency and reduce burden.

**Objective:** To evaluate the reliability of OpenAI’s ChatGPT for performing risk of bias assessments of randomized trials using the revised risk of bias tool for randomized trials (RoB 2.0).

**Methods:** We sampled recently published Cochrane systematic reviews of medical interventions (up to October 2023) that included randomized controlled trials and assessed risk of bias using the Cochrane-endorsed RoB 2.0. From each eligible review, we collected data on the risk of bias assessments for the first three reported outcomes. Using ChatGPT-4, we assessed the risk of bias for the same outcomes using three different prompts: a minimal prompt including limited instructions, a maximal prompt with extensive instructions, and an optimized prompt that was designed to yield the best risk of bias judgements. The agreement between ChatGPT’s assessments and those of Cochrane systematic reviewers was quantified using weighted kappa statistics.

**Results:** We included 34 systematic reviews with 157 unique trials. We found the agreement between ChatGPT and systematic review authors for assessment of overall risk of bias to be 0.16 (95% CI: 0.01 to 0.3) for the maximal ChatGPT prompt, 0.17 (95% CI: 0.02 to 0.32) for the optimized prompt, and 0.11 (95% CI: −0.04 to 0.27) for the minimal prompt. For the optimized prompt, agreement ranged between 0.11 (95% CI: −0.11 to 0.33) to 0.29 (95% CI: 0.14 to 0.44) across risk of bias domains, with the lowest agreement for the deviations from the intended intervention domain and the highest agreement for the missing outcome data domain.

**Conclusion:** Our results suggest that ChatGPT and systematic reviewers only have “slight” to “fair” agreement in risk of bias judgements for randomized trials. ChatGPT-4 cannot be relied upon to judge risk of bias and should not be used for this purpose. Since this study was completed, ChatGPT has advanced and OpenAI has released newer models with different capabilities. These may prove more adept at risk of bias evaluation. There may also be opportunities to use ChatGPT to streamline other aspects of systematic reviews, such as screening of search records or collection of data.

## Background

The practice of evidence-based medicine demands knowledge of the best available evidence, which most often comes from rigorous systematic reviews and meta-analyses (1). Systematic reviews, however, are time- and resource-intensive (2–4). Empirical evidence suggests they typically require upwards of one year to complete and publish and many are outdated at or shortly following publication (3, 5).

One time- and resource-intensive component of systematic reviews is the assessment of risk of bias of primary studies—defined as the propensity for studies to systematically over- or underestimate treatment effects (6, 7). Risk of bias assessments are burdensome and time-consuming and demand specialized training (6, 7). Moreover, to reduce the opportunity for errors, guidance for conducting rigorous systematic reviews typically suggests authors assess risk of bias independently and in duplicate, adding to the complexity and workload of the process (6).

Many tools exist to assess the risk of bias of randomized trials (8, 9), examples of which include the tools from the Joanna Briggs Institute (10), the Jadad Scale (11), and the Critical Appraisal Skills Program (CASP) checklist (12). These tools, however, generally fall short compared to the most commonly used tool, the original Cochrane risk of bias tool for randomized trials, and their application is not recommended (6, 13, 14).

In 2019, a new risk of bias tool was introduced that built on the successes of the previous Cochrane-endorsed risk of bias tool but also incorporated new advancements (15). This tool was called the revised tool for assessing risk of bias of randomized trials (RoB 2.0) and is now largely considered the gold standard (6).

The application of the RoB 2.0 tool, like other risk of bias tools, typically involves reviewers using trial reports and trial registrations or protocols, when available, to make judgements for each risk of bias domain (6, 15). Reviewers who collect data for a systematic review are also typically tasked with assessing the risk of bias of eligible trials (6). The RoB 2.0 tool rates risk of bias as either high, some concerns, or low across five domains: randomization, deviations from intended intervention, missing outcome data, measurement of outcome, and selective reporting. To guide judgements, the RoB 2.0 includes signaling questions for each domain. The overall rating of risk of bias is determined by the domain rated at highest risk of bias (6, 15).

While the RoB 2.0 tool builds off a decade’s worth of experience with the original risk of bias tool, recent evidence suggests that reviewers find it complex and time-consuming—perhaps more complex and time-consuming than previous risk of bias tools (7, 16). Innovations to streamline and simplify risk of bias assessments without compromising their rigor will reduce the time and effort required to perform systematic reviews and aid in maintaining their currency.

RobotReviewer is an automated tool to extract data from and assess the risk of bias of randomized trials (17–19). Previous studies on RobotReviewer show optimistic results, with generally moderate to high agreement with systematic reviewers (70% to 90%) (17, 18). The RobotReviewer, however, was trained on the original Cochrane risk of bias tool, rather than the RoB 2.0 tool, and only offers judgements on four of the seven domains of the original tool. To our knowledge, RobotReviewer is the only artificial intelligence (AI) tool for assessing risk of bias in systematic reviews (20).

ChatGPT (OpenAI, San Francisco, California, USA) is an artificial intelligence language model developed by OpenAI that generates human-like text and engages in conversation by predicting words based on context and training data (21). Differing from specialized automated tools for risk of bias assessments, ChatGPT is was not intentionally created for health research, systematic reviews, or risk of bias assessments (21). ChatGPT has nonetheless demonstrated the capacity to execute complex tasks (22–25) and it may therefore also prove capable of assessing risk of bias. Since its initial release in November 2022, ChatGPT has undergone several iterations, beginning with ChatGPT-3.5, followed by ChatGPT-4 in March 2023, ChatGPT-4o in 2024, and ChatGPT-5 in 2025, which represents the current model. Each successive version has incorporated advancements in reasoning, interactivity, multimodal integration (including file uploads and image generation), transparency, speed, and safety.

This study evaluates the performance of ChatGPT, an AI-based language model, for assessing risk of bias of randomized trials using the RoB 2.0 tool. To do this, we sampled Cochrane systematic reviews using the RoB 2.0 tool and used ChatGPT-4—an advanced large language model offered by OpenAI—to assess the risk of bias of the trials within these reviews. At the time of this study, ChatGPT-4 was the most advanced ChatGPT model publicly available. We compared ChatGPT’s assessment with those presented in Cochrane reviews. Consistency in assessments of risk of bias between ChatGPT and Cochrane reviewers will suggest that ChatGPT can provide a reliable assessment of the risk of bias of randomized trials. Conversely, discrepancies in risk of bias assessments between ChatGPT and Cochrane reviewers will suggest that ChatGPT is unreliable for assessing risk of bias.

We are aware of a concurrent protocol that describes an independent, ongoing study to also evaluate ChatGPT’s performance for assessing risk of bias (26). The protocol informed the reporting and interpretation of the current study. Unlike the current study, the ongoing study is not limited to the RoB 2.0 tool and is expected to evaluate newer iterations of ChatGPT and will therefore yield findings that are different but complementary to those reported here.

## Methods

We registered our protocol on Open Science Framework (https://osf.io/aq85p) in September 2023. We report our study according to the Preferred Reporting Items for Systematic Reviews and Meta-Analyses (PRISMA) checklist (27). A protocol describing an independent, ongoing study to also evaluate ChatGPT’s performance for assessing risk of bias also guided the reporting of this study (26).

This study does not involve human participants and is thus exempt from ethics review.

Figure 1 presents an overview of our methods.

**Figure 1:**
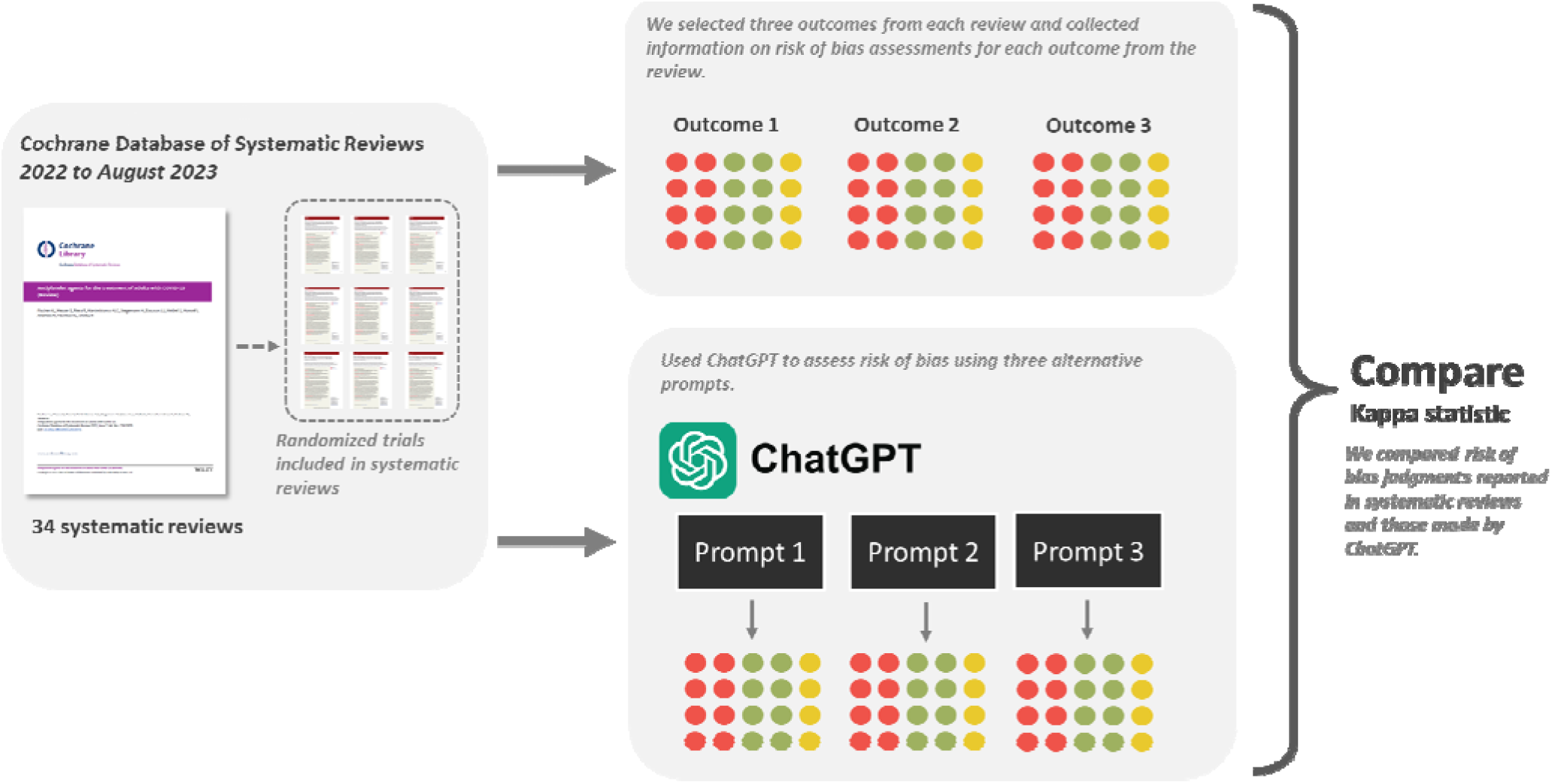
Overview of methods

### Search strategy and screening

We used the Cochrane Database of Systematic Reviews (CDSR), which provides a chronological catalogue of published and updated Cochrane systematic reviews, to identify eligible reviews.

Reviewers worked independently and in duplicate to screen Cochrane reviews for eligibility, starting with the most recently published (August 2023) and working backwards in time. We preferentially included the most recent reviews, as these were more likely to have applied the finalized version of the RoB 2.0 tool rather than earlier pilot versions (15). Screening continued until we identified our target sample size of approximately 160 trials.

### Eligibility criteria

Our sampling approach was designed to include randomized trials addressing a diverse range of questions (i.e., trials that came from different systematic reviews) and both dichotomous and continuous outcomes.

We included newly published or updated Cochrane systematic reviews addressing the benefits and/or harms of health interventions that included one or more parallel randomized trials and reported consensus-based risk of bias judgements using the Cochrane-endorsed RoB 2.0 tool (15). We defined consensus-based as two reviewers agreeing on the final risk of bias judgements. This may involve two reviewers independently assessing risk of bias and resolving conflicts by discussion or a reviewer assessing risk of bias and a second reviewer confirming the first reviewers’ judgements.

We excluded systematic reviews that were not published by Cochrane, since such reviews may not involve reviewers with sufficient training to appropriately apply the RoB 2.0 tool. We also excluded Cochrane systematic reviews that investigated prognosis or the performance of diagnostic tests and systematic reviews that only included observational studies since these reviews will necessitate the use of other risk of bias tools.

Cochrane systematic reviews use summary of findings tables to present their results (6, 28). These tables list outcomes in order of importance, the number of trials and patients that contributed data to the meta-analysis for each outcome, the relative and absolute effect estimates based on meta-analyses, and judgements about the certainty of evidence (6, 28). From each eligible review, we selected the first two listed outcomes (suggesting that they are the most important) that were informed by one or more trials. If either of the first two outcomes were continuous, we then selected the third outcome listed in the summary of findings table. If the two reported outcomes were both dichotomous, we then selected the first listed continuous outcome reported in the summary of findings table. When summary of findings tables reported on the same outcome at different timepoints, we selected entirely unique outcomes.

From each review, we included all parallel randomized trials published in English that were included in analyses addressing the outcomes of interest. We excluded crossover and cluster randomized trials since these trial designs require unique considerations in their assessment of risk of bias and different versions of the RoB 2.0 tool. Cochrane reviews often include unpublished trial data. When reviews reported that information for a particular trial was unpublished or was drawn from a combination of unpublished and published data, we excluded those trials since we did not have access to the same unpublished information as the Cochrane reviewers for risk of bias assessments.

For feasibility, we also excluded trials for which data were drawn from multiple publications. Including such trials would have necessitated an exhaustive review of all related publications to identify those containing the outcome data and the comprehensive details required for risk of bias assessment.

### ChatGPT prompts

A key component in the use of ChatGPT is the design of the text used to instruct the model (called ‘prompts’) to generate an answer. We anticipated that ChatGPT’s risk of bias judgements may depend on the nature of the prompts that it is provided. To study how different prompts may influence risk of bias judgements, we iteratively designed three different prompts: a minimal prompt including limited instructions for assessing risk of bias, a maximal prompt with extensive instructions, and an optimized prompt that was designed to include sufficient information to yield the best risk of bias judgements.

We piloted the prompts using 15 trials drawn from systematic reviews previously performed by our own team and refined the prompts by iterative discussion and input by the co-authors (29–31). All prompts asked ChatGPT to judge risk of bias for all RoB 2.0 domains (bias due to randomization, deviation from intended intervention, missing outcome data, measurement of outcome, and selective reporting) as low risk of bias, some concerns, or high risk of bias—consistent with RoB 2.0 guidance (15). Supplement 1 presents these three prompts.

The RoB 2.0 tool is accompanied by a document that describes the tool and offers guidance on its implementation. All three prompts included the RoB 2.0 full guidance document (riskofbias.info), which were fed to ChatGPT using the AskYourPDF ChatGPT plugin that allows ChatGPT to read and query PDF documents. All prompts also included a PDF copy of the trial publication, a PDF copy of the trial registration or protocol (if one was available), and specified the outcome of interest for which risk of bias assessment was being performed. The prompts also specified the order in which the RoB 2.0 domains should be assessed and that the responses should include a judgment and rationale.

The RoB 2.0 tool offers two options for assessing the risk of bias due to deviations of the intended intervention: one for the effect of assignment to the intervention and the other for the effect of adherence. In Cochrane systematic reviews, the subsection on risk of bias typically reports which option reviewers applied. Our ChatGPT prompts also specified whether to assess risk of bias for the effect of assignment or adherence to the intervention, using the same option used by the Cochrane systematic reviews. For systematic reviews that failed to clarify their appraoch, we assumed they assessed risk of bias for assignment to the intervention.

The ChatGPT prompts do not include any information related to the consensus-based risk of bias judgements presented in the systematic reviews. Hence, ChatGPT is ‘blind’ to the Cochrane systematic reviewers’ risk of bias judgements.

### Data collection

RoB 2.0 guidance demands that reviewers perform risk of bias judgements for each particular result rather than each trial or outcome, since risk of bias may differ across outcomes in a trial or across different ways of statistically summarizing the results for the same outcome (15). We took this approach in this study.

For each eligible trial and outcome, we collected information on the consensus-based risk of bias judgements presented in the Cochrane systematic reviews. Subsequently, for each eligible trial, we used the ChatGPT-4 chatbot to assess the risk of bias of the outcomes of interest, using each of the three ChatGPT prompts. In July and August 2023, when data collection occurred, ChatGPT-4 stood at the most advanced iteration ChatGPT. Unlike its predecessor ChatGPT-3, ChatGPT-4 was only available with a paid subscription to OpenAI.

We implemented each of the prompts in unique chats. We did not collect data in duplicate because the nature of the data did not require any subjective judgements and we anticipated that the only potential source of error is mistakes in copying and pasting prompts to the ChatGPT interface, which we deemed unlikely.

We anticipated that the reliability of ChatGPT may depend on the objectivity of the outcome for which risk of bias is being assessed. We considered outcomes objective if they were based on established laboratory measures or if they were not subject to interpretation by patients or healthcare providers. Conversely, we considered outcomes subjective if they were patient-reported or subject to interpretation by patients or healthcare providers. We classified outcomes as either objective (e.g., mortality), probably objective (e.g., unscheduled physician visits), probably subjective (e.g., serious adverse events), and definitely subjective (e.g., quality of life) to facilitate stratified analyses based on the degree of objectivity of the outcome.

#### Data synthesis and analysis <u>Sample size estimation</u>

We used the kappaSize package in R (Vienna, Austria, Version 4.1.3) to estimate sample size (32). We aimed to calculate the number of required trials to obtain a sufficiently precise estimate of a value of kappa for which systematic reviewers will feel confident using ChatGPT for risk of bias assessments. We assumed that most reviewers would feel confident using ChatGPT if it yields a kappa of 0.70, indicating substantial agreement, with the lower bound of the confidence interval no less than 0.55. We anticipated the risk of bias distribution to be approximately 30% low, 30% with some concerns, and 40% high.

We inflated the estimated sample size by a design effect to account for correlation between the risk of bias of trials from the same review. We assumed an intra-review correlation of 0.05 and an average of 10 trials per review, yielding a design effect of 1.45. This resulted in a minimum sample size of 120 trials from 12 reviews. We investigated the sensitivity of our estimated sample size to different assumptions about the anticipated distribution of risk of bias judgements and the potential correlation between trials from the same review. To account for other potential scenarios (e.g., kappa = 0.6, intrareview correlation of 0.1), we ultimately intended to include approximately 160 trials from 16 reviews.

#### Agreement between ChatGPT and consensus-based risk of bias assessments

We present the inter-rater agreement, represented by weighted kappa, between each of the three ChatGPT prompts and consensus-based risk of bias judgements from Cochrane authors. Unlike percentage agreement, the weighted kappa accounts for the possibility of agreement due to chance and for the ordinal nature of the response options of the RoB 2.0 tool (low risk of bias, some concerns, high risk of bias) (33).

We present separate analyses for each RoB 2.0 domain and for the overall rating of risk of bias. Each analysis only includes one outcome from each included trial. Our primary analysis includes the most important outcome, based on the order in which outcomes were listed in Cochrane systematic review summary of findings tables. We adjusted for clustering of trials within each systematic review by inflating the variance of all estimates by the design effect (34).

We interpreted Cohen’s kappa statistics using the most commonly used thresholds: values from 0.0 to 0.2 indicating slight agreement, 0.21 to 0.40 indicating fair agreement, 0.41 to 0.60 indicating moderate agreement, 0.61 to 0.80 indicating substantial agreement, and 0.81 to 1.0 indicating perfect agreement (35, 36). We acknowledge, however, that these thresholds are not rigid standards and that alternative cutoffs may be appropriate in different contexts.

We hypothesized that ChatGPT may be more reliable to assess risk of bias when there are few subjective judgements. Therefore, we expected better agreement for: (i) trials addressing pharmacologic interventions because trials of pharmacologic interventions are more likely to blind patients and healthcare providers thus simplifying judgements related to deviations from intended intervention and measurement of outcomes; (ii) trials addressing risk of bias of assignment of the intervention because assignment to the intervention does not necessitate making judgements about adherence; (iii) objective outcomes since these outcomes do not need additional judgements about whether failure to blind may have resulted in differential measurement of the outcome; and (iv) dichotomous instead of continuous outcomes since continuous outcomes are more likely to be subjective.

To test these hypotheses, we performed secondary analyses stratified by these factors. We also performed a secondary analysis in which we collapsed ratings of “some concerns” and “high risk of bias” into a single category.

We performed all statistical analyses using the psych package in R (Vienna, Austria, Version 4.1.3) (37).

#### Review of ChatGPT justifications for discrepant risk of bias judgements between Cochrane systematic reviewers and ChatGPT

Our prompts queried ChatGPT to provide a justification for its ratings of risk of bias. To understand reasons why ChatGPT may produce unreliable risk of bias judgements, we also qualitatively reviewed justifications provided by ChatGPT to support its judgements for potential errors or problems.

## Results

### Systematic review and trial characteristics

We included 157 trials from 34 systematic reviews. Figure 2 presents the selection of systematic reviews. Supplement 2 presents a list of included reviews and supplement 3 presents a list of excluded reviews.

**Figure 2:**
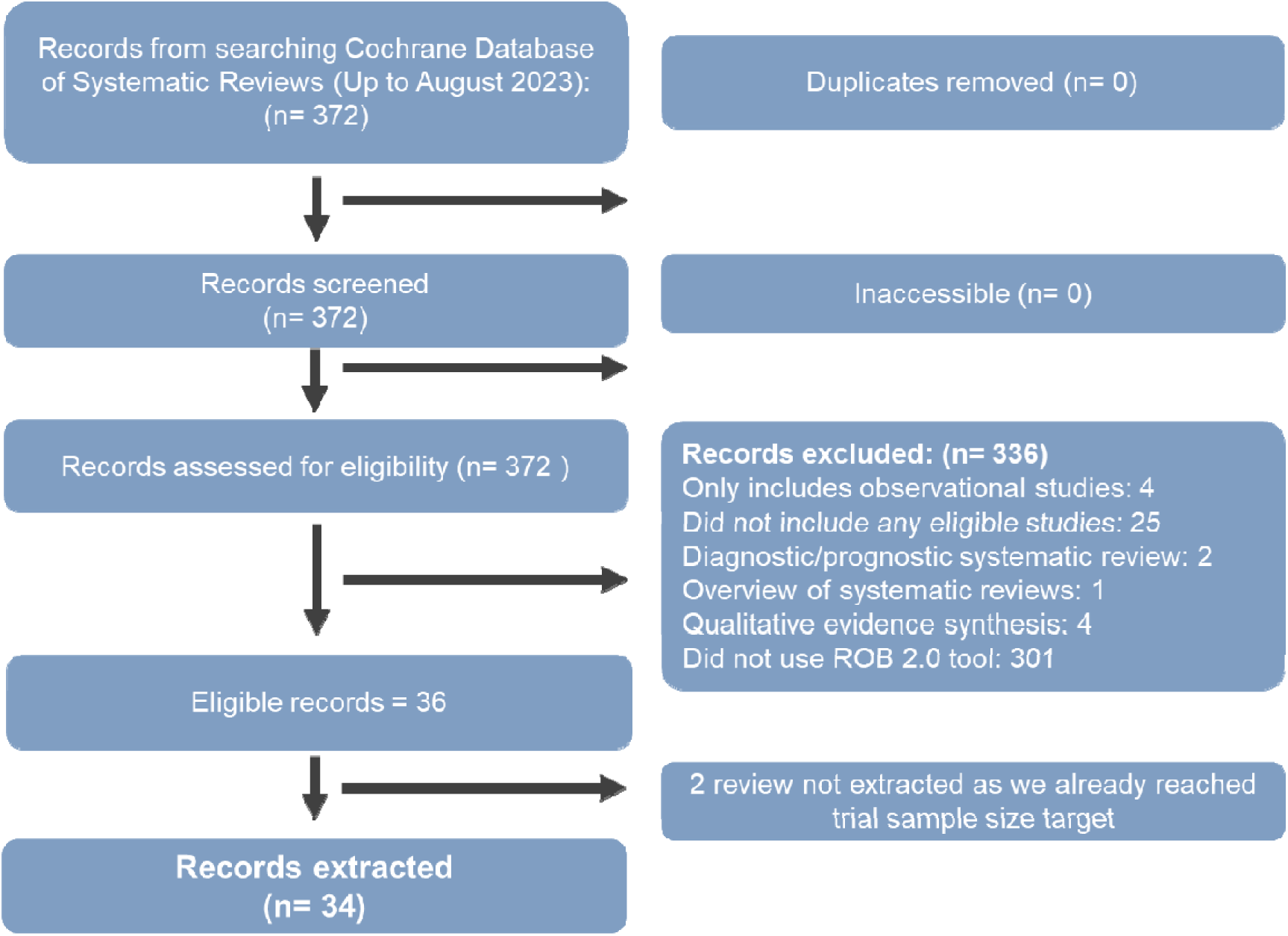
Screening process

**Figure 3:**
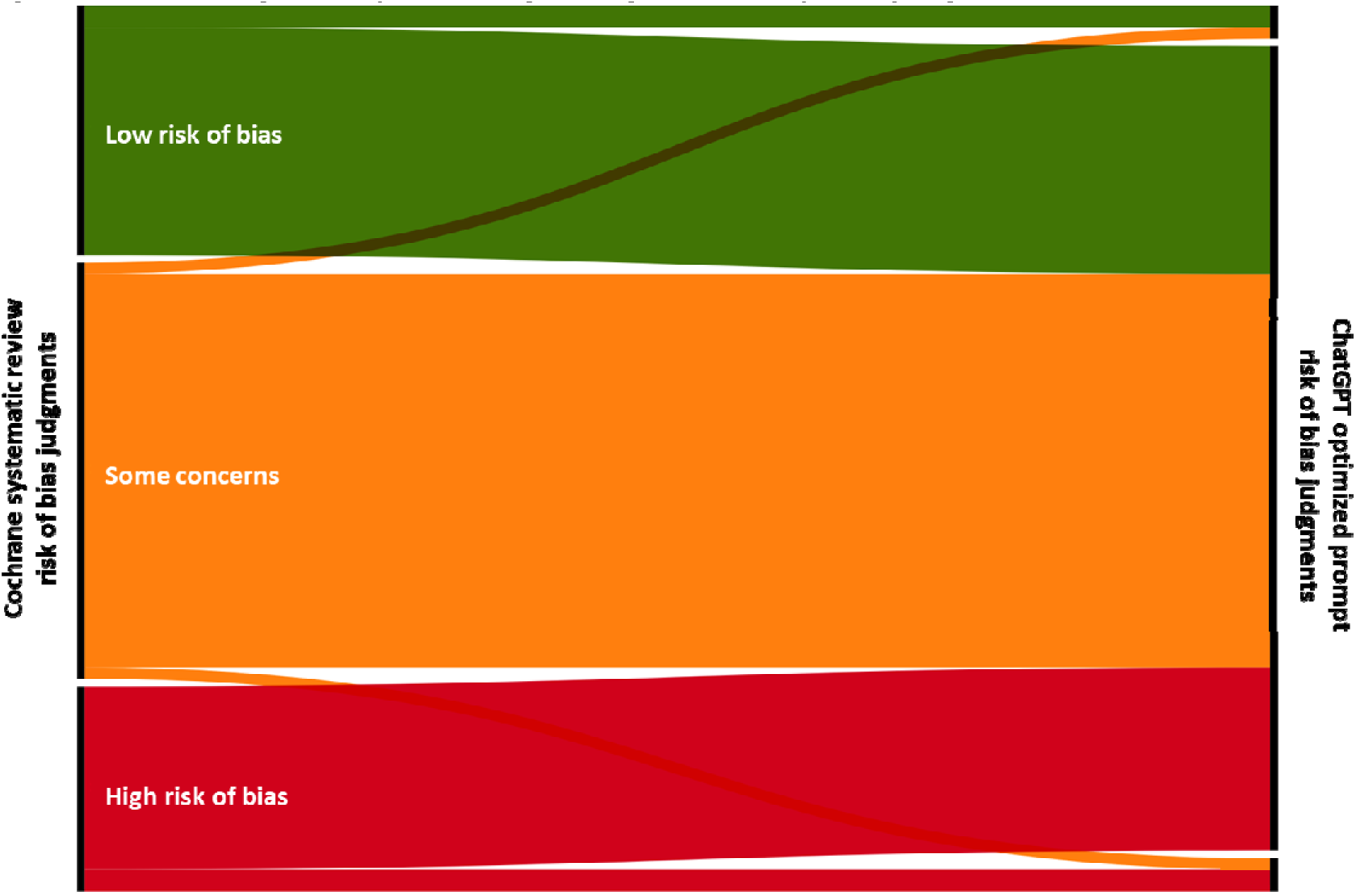
Flow diagram representing changes in risk of bias judgments The bars on the left represent ratings of low risk of bias (represented in green), some concerns (represented in orange), and high risk of bias (represented in red) by Cochrane systematic reviewers, respectively. The bars on the right represent ratings of low risk of bias, some concerns, and high risk of bias by ChatGPT. The graph represents differences in overall risk of bias ratings between Cochrane systematic reviewers and ChatGPT.

More than half of reviews were published in 2023 and addressed pharmacologic interventions. Reviews most addressed infectious, ophthalmologic, and respiratory conditions. Reviews either rated the risk of bias for assignment to the intervention or did not report whether they assessed the risk of bias of assignment to or adherence to the intervention. More than half of included outcomes were dichotomous and rated as either definitely or probably objective.

In our analyses, each trial contributed data only for one outcome. Our primary analysis included data from 157 trials. Of these, 45 (28.7%) were rated at low risk of bias overall by Cochrane systematic reviewers, 75 (47.8%) at some concerns, and 37 (24.6%) at high risk of bias. Fifty-two trials (33.1%) were rated at high risk of bias or some concerns for bias due to randomization, 37 (23.6%) for bias due to deviations from the intended intervention, 23 (14.7%) for missing outcome data, 29 (18.5%) for measurement of the outcome, and 72 (45.9%) for selective reporting.

### Agreement between ChatGPT and consensus-based risk of bias judgements from Cochrane review authors

In our analyses, each trial contributed data only for one outcome. When a trial reported data on more than one outcome of interest, we included data for the outcome reported first in the systematic review.

We found overall only slight agreement between ChatGPT risk of bias judgements and consensus-based risk of bias judgements from systematic reviewers. Agreement for overall risk of bias ranged between 0.11 (95% CI: −0.04 to 0.27) and 0.17 (95% CI: 0.02 to 0.32) for the minimal and optimized prompts, respectively. Figure 2 presents a flow diagram representing categorical changes in the overall rating of risk of bias between systematic reviewers and the optimized ChatGPT prompt.

For the optimized prompt, agreement ranged between 0.11 (95% CI: −0.11 to 0.33) to 0.29 (95% CI: 0.14 to 0.44) across risk of bias domains, with the lowest agreement for the deviations from the intended intervention domain and the highest agreement for the missing outcome data domain.

We did not find evidence that ChatGPT had importantly different reliability in stratified analyses based on whether trials addressed pharmacologic or non-pharmacologic interventions, objective or subjective outcomes, dichotomous or continuous outcomes, or whether reviews specified assessing the risk of bias of assignment to the intervention (Supplements 4 to 10). ChatGPT showed “slight” to “fair” agreement for these subgroups.

Likewise, our secondary analysis that collapsed ratings of “some concerns” and “high risk of bias” into a single category also showed “slight” to “fair” agreement (Supplement 11).

##Supplement 12 presents qualitative observations about divergent risk of bias judgements between ChatGPT and Cochrane systematic reviewers. We identified four problems. First, ChatGPT struggled to distinguish between features signaling low and high risk, often misinterpreting randomization methods or imputing missing data without evidence. Second, it could not make reasonable assumptions where trials required informed inference. Third, it showed unfamiliarity with recommended assessment processes, at times conflating domains such as deviations from intended interventions and outcome measurement. Finally, it committed random errors, leading to inconsistent ratings even within the same trial.

## Discussion

### Main findings

We performed a study evaluating OpenAI’s ChatGPT-4 for assessing the risk of bias of randomized trials using the Cochrane-endorsed RoB 2.0 tool (15). To do this, we sampled Cochrane systematic reviews that reported RoB 2.0 judgements for randomized trials, assessed the risk of bias of trials using ChatGPT via three variations of prompts, and compared the degree of agreement between RoB 2.0 judgements presented in systematic reviews and those made by ChatGPT.

We found only slight to fair agreement between ChatGPT risk of bias judgements and those presented in systematic reviews. Our results suggest that ChatGPT-4 is suboptimal for facilitating risk of bias assessments. We found similar results when we restricted our analysis to subgroups for which we hypothesized that ChatGPT may be more reliable, including trials addressing pharmacologic interventions, reviews assessing the risk of bias associated with assignment to the intervention, objective outcomes, and dichotomous outcomes.

We also reviewed cases in which ChatGPT’s risk of bias judgements differed from those of Cochrane systematic reviewers with the goal of identifying ways in which we can refine future prompts. Our findings indicate that ChatGPT might make more accurate risk of bias judgements if informed about both low and high risk of bias methodological traits. For example, one trial reported randomization by an “interactive web-response system”, which suggests central randomization and allocation concealment (38). ChatGPT, however, rated the trial at some concerns for randomization because the trial report “does not explicitly mention whether the allocation sequence was concealed”. Training ChatGPT to recognize features of trials at low versus high risk of bias may improve the reliability of its risk of bias assessments.

Though our results appear discouraging, they must also be contextualized considering general poor agreement between even experienced reviewers in implementing the RoB 2.0 tool. For example, a previous investigation of the reliability of RoB 2.0 using experienced systematic reviewers reported inter-rater reliability ranging between 0.04 to 0.45, indicating only slight to fair agreement (16). The original Cochrane risk of bias tool also demonstrated poor inter-rater reliability for select domains (39).

Finally, while we evaluated the degree of agreement between risk of bias judgements reported in systematic reviews and those made by ChatGPT, we did not consider the impact of these discrepancies. For example, discrepancies in risk of bias judgements may not necessarily lead to an overall change in the rating of the certainty (quality) of evidence and the material conclusions of systematic reviews.

Strengths and limitation

The primary strength of our study is its generalizability to diverse research questions, reviews, and research teams. Risk of bias judgements are subjective and different research groups and teams may apply distinct thresholds or interpretive lenses when expressing concern. Similarly, the assessment itself invariably reflects nuances specific to the research question under consideration. As our sample included systematic reviews from multiple diverse research teams, ChatGPT’s reliability is not confined to the specific idiosyncrasies of a single group’s approach to risk of bias assessments or to a single topic.

Our study was limited to parallel randomized trials published in English. We excluded crossover and cluster randomized trials since these trial designs require unique considerations in their assessment of risk of bias and different versions of the RoB 2.0 tool. Thus, the results of our study may lack generalizability beyond English language parallel randomized trials, though these are the most common studies typically included in systematic reviews. Further, it is unlikely for ChatGPT to be able to perform remarkably differently for other types of trials, since assessing the risk of bias of these trials necessitates the same considerations as parallel randomized trials in addition to several other unique considerations.

Evidence suggests that risk of bias assessments in Cochrane reviews, despite their rigor, are sometimes unreliable and inconsistent with established guidance (16). Hence, differences in risk of bias judgements between ChatGPT and Cochrane systematic reviewers may also represent errors on part of reviewers. Previous studies suggest that agreement between reviewers in assessing risk of bias may be very poor (40, 41). To minimize the potential for this error, we limited our sample to Cochrane systematic reviews, which are known for their methodological rigor (42, 43).

ChatGPT’s performance is also not static. The infrastructure, interfaces, and applications built around ChatGPT are continuously updated (44–46). Our experiment relied on ChatGPT-4, the most advanced version available in 2023 but now retired. It is likely that the performance that we observed will not replicable in the future—though it is more likely that the capabilities of ChatGPT have improved rather than deteriorate. Even with identical prompts, ChatGPT might provide slightly different answers due to the inherent stochasticity in its response generation (45).

The reliability of ChatGPT risk of bias assessments is likely to depend on the nature of the prompts. We tested three different prompts. Our results suggest that the performance of the three prompts is comparable. It is possible that reviewers may be able to produce more reliable risk of bias assessments using alternative prompts.

Our prompts queried ChatGPT to provide a justification for its ratings of risk of bias. To understand reasons why ChatGPT may produce unreliable risk of bias judgements, we also reviewed justifications provided by ChatGPT to support its judgements for potential errors or problems. While we performed a general review of justifications for which ChatGPT and Cochrane reviewers made discrepant risk of bias judgements, we did not perform a formal qualitative analysis of the justifications.

While we did not record the exact duration our team spent using ChatGPT, we estimate that each trial took no longer than 15 minutes—much less time than on average required for a reviewer to conduct an individual risk of bias assessment and consensus meeting according to empirical evidence (7, 16).

Finally, our systematic review includes minor deviations from the protocol. To account for correlation between trials in the same systematic review, we planned to calculate weighted kappa within each review individually and pool the weighted kappa statistics across systematic reviews using random-effects meta-analysis (47). The sampling distribution of kappa, however, is asymmetric. While with a large enough number of observations, the sampling distribution of kappa is approximately normal, we found there to be too few trials within each systematic review to assume normality, precluding our approach to perform meta-analyses. Instead, we adjusted the variance of all estimates for the correlation within each systematic review. Likewise, in our primary analysis, we excluded ratings of uncertain risk of bias from analyses. We had planned to perform additional sensitivity analyses treating these ratings as some concerns or high risk of bias but there were too few uncertain ratings to affect estimates of reliability.

### Relation to previous research

Attempts to reduce the time, resources, and expertise needed to perform systematic reviews are not new. For example, RobotReviewer is an automated tool to extract data from and assess the risk of bias of randomized trials (17). The RobotReviewer, however, was trained on the original Cochrane risk of bias tool and only offers judgements on four of the seven domains of the original tool. Since then, Cochrane has adopted a revised risk of bias assessment tool that requires more nuanced judgements and is more resource- and time-intensive (7). Given the performance of ChatGPT, however, adapting RobotReviewer to provide risk of bias assessments using the RoB 2.0 tool may be more promising.

Since the completion of the current study, several new studies have also reported similar investigations about the utility of ChatGPT for performing risk of bias assessments using newer ChatGPT models, one focusing on the CLARITY risk of bias tool and another on neonatology trials (48–50). Although findings were mixed, the collective evidence continues to indicate that ChatGPT is not reliable for fully automated risk of bias assessments. Notably, the study applying the CLARITY risk of bias tool found ChatGPT to perform more consistently (50).

### Implications

Our results suggest that ChatGPT, in the form in which it was tested in this study (ChatGPT-4), is not able to reliably assess the risk of bias of randomized trials. Since assessment of the risk of bias of observational and diagnostic studies is even more complicated, it is reasonable to expect that ChatGPT might encounter even more challenges with these other types of study designs.

Since we completed this study, ChatGPT has advanced considerably, and OpenAI has introduced newer models with distinct capabilities. Our experiment relied on ChatGPT-4, the most advanced version available in 2023 but now retired. In November 2023, OpenAI released the option to create custom GPTs; in 2024, it released GPT-4o and several variants; and in 2025, GPT-5 and its variants. These advances represent material progress in reasoning, memory, context window, tools, plugins, and customization—developments that will likely have substantial implications for ChatGPT’s capacity to conduct risk of bias assessments. The results we observed in this study cannot be extrapolated to newer OpenAI models.

Our study also has implications for future research. The prompts we designed could not yet assess risk of bias with reliability, but alternative prompts may yield more dependable results. Each domain of the RoB 2.0 tool, for example, is structured around signaling questions that guide reviewers to evaluate aspects of trial conduct that may introduce bias. These questions require structured responses—“Yes,” “Probably yes,” “Probably no,” “No,” or “No information”—which are then synthesized into a judgment of “Low,” “Some concerns,” or “High” risk of bias. Rather than asking ChatGPT to deliver domain-level judgments directly, future work might prompt it to engage systematically with these signaling questions.

Likewise, ChatGPT may be prompted to consider each domain separately, which, due to limitations in its memory, may improve its performance. With the recent ability to work across files and projects, supplying ChatGPT with risk of bias manuals may further enhance its accuracy. Future research may also address the usefulness of having systematic reviewers reconcile their risk of bias assessments with ChatGPT or the role of ChatGPT in training systematic reviewers.

There are also opportunities to use ChatGPT to streamline other aspects of systematic reviews. Early studies suggest that ChatGPT can be used to devise search strategies (24). ChatGPT may also assist with screening search records, extracting data from eligible studies, or performing evaluations of the certainty of evidence. Screening studies is less subjective and perhaps better suited to ChatGPT’s abilities.

We are aware of a concurrent protocol that describes an independent, ongoing study to evaluate ChatGPT’s performance for assessing risk of bias, which informed the design and reporting of the current study (26). This protocol was published before publication of our preprint; we became aware near the end of data collection and subsequently acknowledge its influence on our interpretation and reporting. We anticipate this ongoing study will focus on the currently available ChatGPT model. The study appears comprehensive and rigorous in its design and will be able to clarify whether newer OpenAI models can provide more reliable risk of bias assessments.

If research shows that new ChatGPT models may reliably perform risk of bias assessments or if other tools emerge that can more reliably perform various systematic review tasks, systematic review authors will need to consider whether the time and resource savings afforded by these tools are worth potential suboptimal performance. While these tools may not always perform perfectly, they may still be useful in situations in which systematic reviews need to be performed quickly or with limited resources. Similarly, systematic review authors will also need to consider the acceptability of such tools by evidence users. For example, evidence users may be skeptical of systematic reviews that rely on AI tools.

The integration of artificial intelligence and large language models in systematic reviews can also affect trust in health research. We anticipate that due to limited experience, evidence users will be more cautious about the application of studies that use such tools (51, 52).

## Conclusion

We performed a study evaluating the usefulness of ChatGPT for assessing the risk of bias of parallel randomized trials using the Cochrane-endorsed RoB 2.0 tool. We found only slight to fair agreement between ChatGPT risk of bias judgements and risk of bias judgements presented in systematic reviews. Our results suggest that OpenAI’s ChatGPT-4 is suboptimal for performing risk of bias assessments.

Since this study was completed, ChatGPT has advanced and OpenAI has released newer models with more advanced capabilities, which may prove more reliable at risk of bias assessments. The practice of evidence-based medicine demands knowledge of the best available evidence, which most often comes from rigorous systematic reviews. Systematic reviews, though, are time and resource intensive. Tools to assist with systematic reviews, be it with risk of bias assessments or other tasks, are critically needed.

**Table 1:**
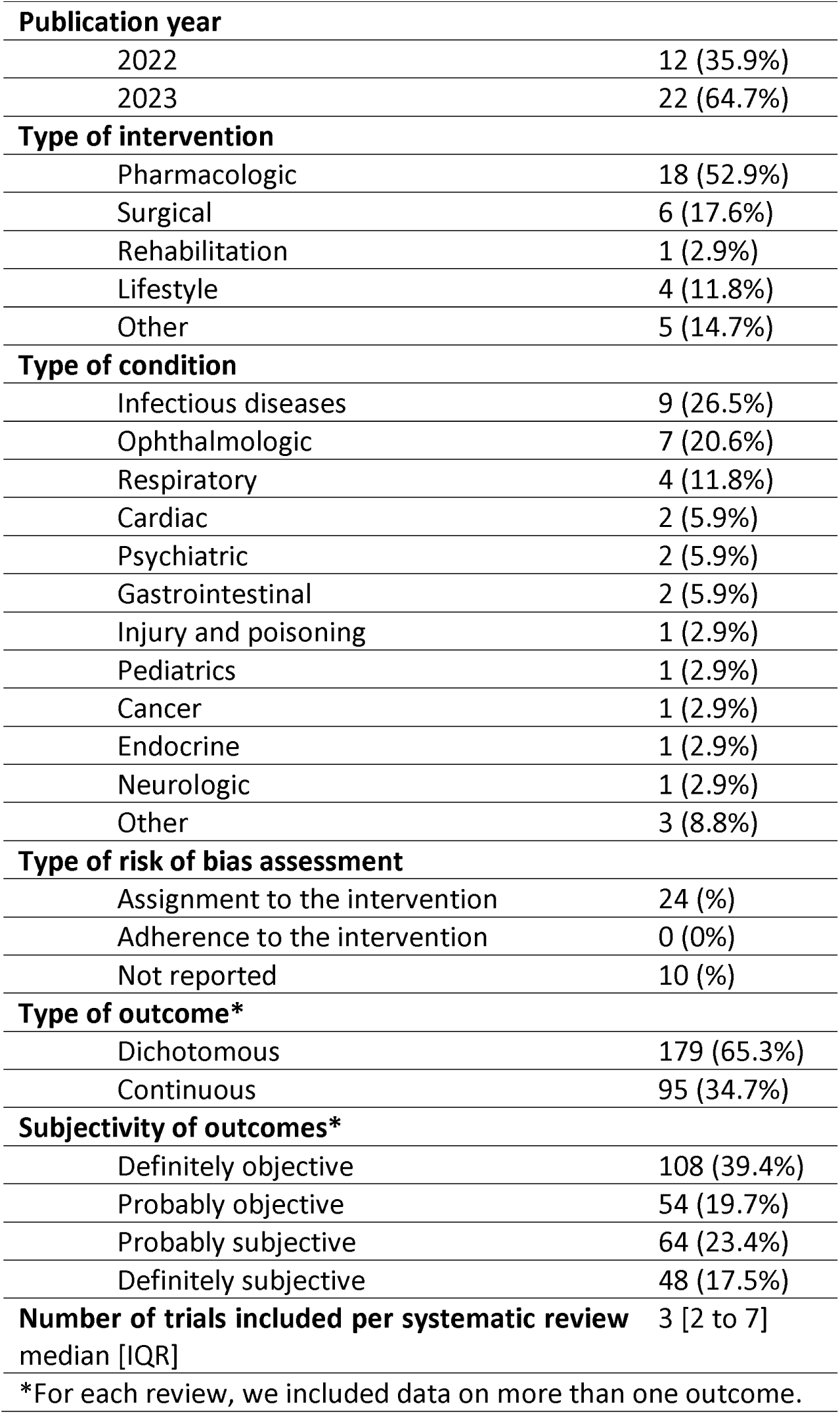
Characteristics of included systematic reviews.

**Table 2:** Degree of Agreement.

|  |  | Consensus based risk of bias judgments reported in systematic reviews |  |  |
| --- | --- | --- | --- | --- |
|  |  | Low risk of bias | Some concerns | High risk of bias |
| **Optimized ChatGPT prompt** | **Low risk of bias** | 4 (2.55%) | 2 (1.27%) | 0 (0%) |
|  | **Some concerns** | 41 (26.11%) | 71 (45.22%) | 33 (21.02%) |
|  | **High risk of bias** | 0 (0%) | 2 (1.27%) | 4 (2.55%) |
| **Minimal ChatGPT prompt** | **Low risk of bias** | 3 (1.91%) | 5 (3.18%) | 1 (0.64%) |
|  | **Some concerns** | 42 (26.75%) | 66 (42.04%) | 32 (20.38%) |
|  | **High risk of bias** | 0 (0%) | 3 (1.91%) | 4 (2.55%) |
| **Maximal ChatGPT prompt** | **Low risk of bias** | 1 (0.64%) | 2 (1.27%) | 0 (0%) |
|  | **Some concerns** | 44 (28.03%) | 72 (45.86%) | 31 (19.75%) |
|  | **High risk of bias** | 0 (0%) | 1 (0.64%) | 6 (3.82%) |

**Table 3:** Weighted kappa values representing the degree of agreement between ChatGPT prompts and systematic review risk of bias judgments.

|  | **Optimized prompt** | **Minimal prompt** | **Maximal prompt** |
| --- | --- | --- | --- |
|  | **Weighted kappa (95% CI)** |  |  |
| Overall risk of bias rating | 0.17 (0.02, 0.32) | 0.11 (-0.04, 0.27) | 0.16 (0.01, 0.3) |
| Risk of bias due to randomization | 0.24 (0.02, 0.47) | 0.09 (-0.16, 0.33) | 0.09 (-0.15, 0.34) |
| Risk of bias due to deviations from the intended intervention | 0.11 (-0.11, 0.33) | 0.12 (-0.12, 0.37) | 0.12 (-0.13, 0.36) |
| Risk of bias due to missing outcome data | 0.29 (0.14, 0.44) | 0.23 (0.02, 0.45) | 0.16 (-0.05, 0.36) |
| Risk of bias due to measurement of the outcome | 0.14 (-0.13, 0.41) | 0.04 (-0.18, 0.25) | 0.05 (-0.18, 0.28) |
| Risk of bias due to selective reporting | 0.17 (-0.03, 0.37) | 0.29 (0.08, 0.49) | 0.21 (0.04, 0.37) |

## Disclaimers

None.

## Funding

None.

## Data

Available on OSF.

## Data Availability

Data: Available on OSF

https://osf.io/aq85p

## Acknowledgements

We acknowledge the concurrent work by Dr. Chris Rose and colleagues, whose protocol describing a study on evaluating ChatGPT for risk of bias assessments was publicly available near the completion of our data collection. It informed our reporting and interpretation—in particular, how we contextualize large-language-model (LLM) outputs within systematic-review workflows—but did not alter our sampling, eligibility, prompts, analyses, or results.

## Authors’ Contributions

DZ and TP conceived this study. TJ, JRT, MS, and ML collected data. DZ and TP analyzed the data. DZ and TP wrote the first draft of the manuscript and all authors reviewed and approved the final version.

## Notes

### Competing Interest Statement

The authors have declared no competing interest.

### Summary of Updates

We submit a revision to correct lack of citations and to correct previous identified errors.

